# Effect of a Third COVID-19 Vaccine Dose on the Incidence of Long COVID Among Adults Who Completed a Primary Vaccine Series: a Target Trial Emulation in a Community-Based Cohort

**DOI:** 10.1101/2025.11.03.25339444

**Authors:** Yanhan Shen, Zach Shahn, McKaylee M. Robertson, Kelly Gebo, Denis Nash, the CHASING COVID Cohort Study Team

## Abstract

**Background:** Evidence on whether a third COVID-19 vaccine dose lowers long COVID risk is mixed. We estimated the effect of receiving ≥1 third dose versus completing only a primary series on 6- and 12-month long COVID incidence using a target-trial emulation in a U.S. community cohort.

**Methods:** We analyzed the CHASING COVID Cohort, a prospective, community-based study of U.S. adults. Eligible participants were ≥18 years, had completed a two-dose primary series, had no prior long COVID, and had no SARS-CoV-2 infection in the 3 months before time zero. Strategies compared were: receive a third dose at time zero vs. not receive a third dose during follow-up. Long COVID was defined as ≥1 new symptom at or beyond 3 months post-infection with concurrent activity limitation, both absent in the prior year. Follow-up was 6 and 12 months. We used a per-protocol analog: participants were artificially censored upon deviating from their assigned strategy or lost-to-follow-up, with inverse-probability weights to address selection due to censoring and time-varying confounding. We fit weighted pooled logistic models to estimate weighted incidence, differences, and ratios at each horizon.

**Results:** Across 16 sequential trials (18,930 person-trials; 4,044 unique individuals), 3,321 person-trials received a third dose at time zero and 15,609 did not. At 6 months, weighted long COVID incidence was 0.9% (95% CI, 0.5%, 1.3%) with a third dose vs. 1.0% (0.8%, 1.1%) without (risk difference (RD), −0.1%; 95% CI, −0.5%, 0.4%; risk ratio (RR), 0.93; 95% CI, 0.54, 1.44). At 12 months, incidence was 4.9% (4.1%, 5.9%) with a third dose vs. 4.5% (4.1%, 4.8%) without (RD, 0.4%; 95% CI, −0.5%, 1.4%; RR, 1.09; 95% CI, 0.90, 1.33).

**Conclusion:** In this community-based target-trial emulation, receiving a third COVID-19 vaccine dose did not meaningfully reduce 6- or 12-month long COVID incidence compared with completing only a primary series.

## INTRODUCTION

A portion of individuals continue to experience symptoms three months following an acute COVID-19 infection that they had not experienced prior to infection, a condition collectively termed long COVID, which the National Academies describe as an infection-associated chronic state with persistent, relapsing, or progressive symptoms that often lead to activity limitations— such as impairments in daily home functioning, employment, or education ^1–3^. Based on a nationally representative sample of U.S. adults as of December 2023, 8.4% (95% CI, 8.0%-8.8%) of adults reported they had ever experienced long COVID, 3.6% (95% CI, 3.3%-3.9%) reported they currently had long COVID, and 2.3% (95% CI, 2.1%-2.5%) reported they currently had activity-limiting long COVID, regardless of their COVID-19 infection history ^4^.

Staying up to date on COVID-19 vaccines could prevent or reduce the burden of long COVID, either by preventing infection ^5–10^ or by preventing or mitigating long COVID symptoms, given an infection ^11,12^. The findings across studies have been mixed. Several studies found that completion of the third dose after completing the primary series was associated with diminished risk for long COVID, when compared to receipt of primary series alone or no vaccination at all ^13–19^. In a Spanish cohort study, Domènech-Montoliu et al. found lower long COVID incidence after a third vaccine dose ^13^. Two U.S. cross-sectional studies (Romeiser et al. and Xie et al.) similarly reported reduced odds of long COVID among those who received a third dose compared to partially vaccinated or unvaccinated individuals ^14,15^. In contrast, other studies have found no clear protective effect of booster doses ^20,21^. Ballouz et al. conducted a pooled analysis of two prospective longitudinal cohorts, found that booster vaccination did not consistently reduce the incidence of long COVID across SARS-CoV-2 variants ^20^. In a cross-sectional study, Kahlert et al. similarly found no significant difference between those who received booster doses to those who had received only the primary series or remained unvaccinated ^21^. These mixed findings underscore ongoing uncertainty about whether COVID-19 vaccination beyond the primary series reduces the risk of long COVID.

The reasons for mixed results on the role of COVID vaccines in preventing or reducing the burden of long COVID are likely multifactorial, including difference in study design, population characteristics, evolving immunity, circulating variants, availability of treatments (e.g., monoclonal antibodies, Paxlovid), and varying definitions of long COVID across studies. First, many studies employed cross-sectional designs, which are prone to selection biases, as individuals with persistent symptoms might be more motivated to participate, potentially inflating prevalence estimates. In addition, because data were captured at a single point in time, these studies were unable to assess incidence, limiting the feasibility of causal inference. Second, most existing studies have relied on electronic health records (EHR) or claims data, which are limited to individuals who have accessed healthcare and may not capture the broader community, thereby limiting generalizability. Third, relatively few studies adopted target trial emulation (TTE) designs, which are a recommended approach for strengthening causal inference in observational studies and mitigate biases like immortal time bias. These gaps underscore the need for methodologically rigorous studies of community-based populations to establish clearer causal relationships. To address this, we used a community-based prospective cohort not restricted to healthcare system enrollees and applied a TTE framework to estimate the impact of receiving at least a third (v.s. no third dose) COVID-19 vaccine on the incidence of 6- and 12-month long COVID among participants who completed a two-dose primary series vaccine.

## METHODS

### Participants

The Communities, Households, and SARS-CoV-2 Epidemiology (CHASING) COVID Cohort study, launched on March 28, 2020, was a national, community-based prospective cohort study of adults residing in the U.S. or its territories ^22^. Through internet-based recruitment methods, the cohort represents a socio-demographically and geographically diverse sample of U.S. adults. Recruitment and follow-up procedures have been detailed in previous publications ^23–25^. Between March 2020 and December 2023, participants completed approximately quarterly online assessments covering a range of topics, including health insurance coverage, food insecurity, housing instability, SARS-CoV-2 infection, long COVID symptoms, activity limitations, and COVID-19 vaccination receipt. To detect infection-induced seroconversion, dried blood spot (DBS) samples were collected and tested in four rounds between 2020 and 2023 for total antibodies to the SARS-CoV-2 nucleocapsid protein using the Bio-Rad Platelia assay, which detects IgA, IgM, and IgG, with manufacturer-reported sensitivity of 98.0% and specificity of 99.3% ^26–28^. The study was approved by the Institutional Review Board of the City University of New York (CUNY).

### Exposure: the Third Dose of COVID-19 Vaccine

Participants reported detailed information on their COVID-19 vaccinations across 13 follow-up assessments conducted between December 2020 and December 2023, including the number of doses received, vaccination dates for each dose, and the vaccine manufacturer. For this analysis, we focused on individuals who completed both doses of two-dose primary vaccine series. This included vaccines approved to be administered in the U.S.: Pfizer-BioNTech, Moderna, and Novavax ^29–31^. Participants who received the Johnson & Johnson (J&J) vaccine were excluded, as only about 5% of participants received J&J and its single-dose mechanism complicates interpretation of third-dose vaccination following a two-dose primary series, which was the focus of this analysis. The date of primary series completion was defined as the date of receipt of the second dose. The exposure of interest in this study was the receipt of a third COVID-19 vaccine dose. This third dose included either the 2021–2022 monovalent booster or the 2022–2023 bivalent booster, administered after the primary series ^32,33^.

### Outcome: Long COVID Status

The primary long COVID case definition was based on case definitions from the World Health Organization (WHO) ^34^, the Centers for Disease Control and Prevention (CDC) ^1,35^, and the National Academies of Science, Engineering, and Medicine (NASEM) ^2^. We defined individuals with long COVID as participants who experienced at least one symptom and concurrent activity limitations between 3 and 12 months post-infection, neither of which was present in the year preceding infection. The long COVID symptoms include fatigue, post-exertional malaise, trouble concentrating/brain fog, dizziness, erratic heartbeat, gastrointestinal issues, and loss or alteration of taste or smell. These symptoms were assessed roughly every three months between November 2020 and December 2023. Activity limitations were assessed concurrently with long COVID symptoms and defined as difficulty performing daily activities or household responsibilities ^36^. The details of infection and infection date assignment was documented in Appendix 1.

### Study design: Sequential Target Trial Emulation Using a Cohort

This study aimed to estimate the effect of receiving at least a third COVID-19 vaccine dose on the 6- and 12-month incidence of long COVID compared to only completing a two-dose primary vaccine series. We employed TTE, a framework for designing and analyzing observational studies to address causal questions when a randomized controlled trial (RCT) is impractical due to ethical, logistical, or resource constraints. The TTE approach attempts to estimate the same effect that would be estimated by a hypotehtical RCT by explicitly specifying elements of the hypothetical RCT, including clear definitions of eligibility criteria, treatment strategies, the alignment of time zero (start of follow-up), and outcomes assessment. We first specified the hypothetical RCT protocol, then implemented the observational emulation of this protocol ^37^. Furthermore, our nested sequential trial design maximizes statistical efficiency and enhances precision by utilizing available longitudinal data. Detailed specifications of the emulated sequential target trial were provided in **Supplementary Table 1**.

#### Eligibility Criteria and Treatment Strategy

Participants were eligible for inclusion if they were aged 18 years or older and had received both doses of a two-dose primary COVID-19 vaccine series (Pfizer-BioNTech, Moderna, or Novavax). Participants were excluded if they had any prior history of long COVID (as defined above), a documented SARS-CoV-2 infection within the three months preceding the start of follow-up (time zero), or had received any additional COVID-19 vaccine doses beyond the two-dose primary series before that trial’s time zero (**Supplementary Figure 1**). Participants were assigned to one of two treatment strategies: receiving at least a third dose of COVID-19 vaccine or not receiving a third dose.

#### Sequential Trials, Time Zero, and Follow Up

To account for and leverage participants meeting eligibility criteria at different time points, a series of sequential target trials were emulated monthly over a 16-month period, from September 2021 (when third doses first became publicly available in the U.S.) through December 2022 ^38^. Time zero for each sequential trial was defined as follows: For exposed individuals, time zero was the date they received their third dose of the COVID-19 vaccine. In the same month, unexposed individuals—those who had not received a third dose by the end of the month—were identified, and the last day of that month was assigned as their time zero. Eligibility for each sequential trial was determined based on time-updated, participant-specific characteristics. The sequential trial design is illustrated in **Supplementary Figure 1&2**.

The primary outcomes were the 6- and 12-month incidence of long COVID, assessed from trial time zero. Participants were followed until the earliest occurrence of one of the following events: meeting criteria for long COVID, loss to follow-up (defined as any missed follow-up assessment), or the end of the designated follow-up period (6 and 12 months from time zero). Once participants were censored, they did not re-enter the sample for that trial.

#### Per-protocol Effects

Randomization of the two exposure groups was emulated by adjusting for confounders of the receipt of the third COVID-19 vaccine dose at time zero and subsequent incidence of long COVID. To further account for evolving SARS-CoV-2 variants and changes in state government guidelines regarding booster dose eligibility, calendar month at time zero was also included and modeled using a natural cubic spline with 4 degrees of freedom to flexibly capture nonlinear time trends.

We assessed the per-protocol (PP) effects as our primary causal contrast of interest. This effect represents the difference in the expected incidence of long COVID under two hypothetical scenarios: one in which all participants had received at least a third dose of the COVID-19 vaccine at time zero, and another in which no participants had received a third dose at any point during follow-up. In this framework, receipt of additional vaccine doses beyond the third was not considered a deviation from the treatment strategy.

### Statistical Analysis

#### Time Zero Characteristics and Covariate Balance

Descriptive statistics for time zero characteristics were presented as mean with standard deviation (SD), median with interquartile range (IQR), or frequencies with proportions (details of these characteristics were provided in Supplementary Files Appendix 2). Standardized mean differences (SMDs) were used to compare characteristics of unique participants who received at least a third dose of COVID-19 vaccine as of December 2022 verse those who only received two doses of primary vaccine, as well as person-trial comparisons between those who received a third dose verse those who did not at time zero. Additionally, weighted SMDs were calculated using inverse probability of treatment weighting (IPTW) combined with inverse probability of censoring weighting (IPCW) under the per-protocol framework to assess covariates balance in the analytic sample. An SMD of 0.1 or greater is often interpreted as indicating a meaningful imbalance in covariates as a rule of thumb ^39–41^.

#### Estimating Per-Protocol (PP) Effects

We employed an IPTW approach to adjust for time-fixed and time-varying confounding (details of these characteristics were provided in Supplementary Files Appendix 2) ^42,43^. We used IPCW to adjust for potential selection bias introduced by artificial censoring. We assumed that factors associated with loss to follow-up or deviation from the treatment arm at the time of censoring included: age, gender, race/ethnicity, education, household income, comorbidities, prior SARS-CoV-2 infection history, obesity status, smoking status, health insurance status, access to a primary care provider, region of residence, food insecurity, housing instability, and calendar month of censoring. Spline terms for age and calendar month were also included in the IPCW models to flexibly capture nonlinear relationships. We estimated the time-varying weights by fitting a pooled logistic model for the monthly probability of remaining uncensored, including variables covariates described above.

The PP analysis, person-time was censored at the earliest occurrence of either deviation from the assigned treatment strategy—defined as receiving a third dose in the control group —or loss to follow-up during the 6- and 12-month follow-up periods. To estimate the effect of a third dose of COVID-19 vaccine on risk of long COVID, we fitted a pooled logistic regression model weighted by the product of IPTW and IPCW. The model included treatment arm and follow-up time, with additional interaction terms between treatment and both linear and quadratic time to allow for time-varying effects. We used the predicted probabilities from this weighted model to estimate discrete hazards and calculate adjusted cumulative incidence under each treatment strategy. Risks were derived from survival probabilities, computed as the cumulative product of one minus the estimated hazard at each time point. From these estimates, we calculated the 6- and 12-month cumulative incidence (risk), risk differences, and risk ratios. Percentile-based 95% confidence intervals (CIs) were obtained using nonparametric bootstrapping with 500 samples. Weighted cumulative incidence curves for each treatment group were also estimated using the same model to visualize changes in risk over time ^44^.

#### Sensitivity Analysis

We conducted the following sensitivity analyses to assess the robustness of our findings. (1) To explore variations in long COVID case definitions, we conducted sensitivity analyses using three alternative definitions (Supplementary Table 2A&B). The first alternative definition required at least one symptom at least 3 months post-infection, which was absent pre-infection. The second alternative definition required at least one symptom to be present at two distinct time points: an initial occurrence 3 months post-infection, followed by a second occurrence at least 60 days later. The symptoms at these two time points were not required to be the same, but both had to be absent in the year preceding the index infection date. The third alternative definition classified cases based on self-identified long COVID, defined as “experiencing symptoms more than 4 weeks after you first had COVID-19 that are not explained by something else?”, based on the U.K.’s Office for National Statistics (ONS) case definition that was collected in each follow-up assessment since June 2022 ^45,46^. These alternative definitions allowed us to assess the sensitivity of our findings under different criteria for long COVID classification. (2) We conducted the sensitivity analysis separately for individuals who were infection naïve to SARS-CoV-2 as of time zero, to account for potential differences in hybrid immunity, prior exposure, and risk of long COVID (Supplementary Table 3). We used SAS version 9.4 (SAS Institute, Cary, NC, USA) for data management and R version 2024.09.0+375 (R Foundation for Statistical Computing, Vienna, Austria) for data analyses.

## RESULTS

A total of 18,930 person-trials were emulated across 16 sequential target trials, with 3,321 person-trials in which participants receive a third COVID-19 vaccine dose at time zero and 15,609 person-trials in which participants did not (Table 1). Before weighting, participants who received a third COVID-19 vaccine dose were more likely to be older, with 12.1% aged 65 or older compared to 8.3% in the group did not receive a third dose (SMD=0.191). In terms of educational attainment, 71.9% of individuals who received a third COVID-19 vaccine dose had completed college or higher education compared to 56.5% of participants who did not receive a third dose (SMD=0.326). Additionally, individuals who received a third COVID-19 vaccine dose were less likely to report food insecurity (15.7% vs. 27.7%, SMD=0.297) compared to their counterparts who did not receive a third dose. After IPTW, these imbalances were markedly reduced: the weighted SMDs for age ≥ 65, education, and food insecurity fell to 0.013, 0.089, and 0.088, respectively.

**Table 1:** Time-zero characteristics of eligible participants among person-trial population by COVID-19 vaccine status across 16 sequential trial, The CHASING COVID Cohort Study, September 2021 – December 2023

|  | Received the third dose | Not received the third dose | SMD * | Weighted SMD |
| --- | --- | --- | --- | --- |
|  | N=3,321 | N=15,609 |  |  |
| **Age category** |  |  |  |  |
| 18-29 | 657 (19.8%) | 3883 (24.9%) | 0.191 | 0.013 |
| 30-39 | 891 (26.8%) | 4590 (29.4%) |  |  |
| 40-49 | 603 (18.2%) | 2878 (18.4%) |  |  |
| 50-64 | 769 (23.2%) | 2956 (18.9%) |  |  |
| >=65 | 401 (12.1%) | 1302 (8.3%) |  |  |
| **Gender** |  |  |  |  |
| Male | 1538 (46.3%) | 6926 (44.4%) | 0.041 | 0.026 |
| Female | 1677 (50.5%) | 8205 (52.6%) |  |  |
| Non-Binary | 106 (3.2%) | 478 (3.1%) |  |  |
| **Race/Ethnicity** |  |  |  |  |
| Hispanic | 462 (13.9%) | 2946 (18.9%) | 0.256 | 0.072 |
| White Non-Hispanic | 2247 (67.7%) | 9053 (58.0%) |  |  |
| Black Non-Hispanic | 236 (7.1%) | 2005 (12.8%) |  |  |
| Asian Non-Hispanic | 290 (8.7%) | 1180 (7.6%) |  |  |
| Others | 86 (2.6%) | 425 (2.7%) |  |  |
| **Education** |  |  |  |  |
| No College | 932 (28.1%) | 6786 (43.5%) | 0.326 | 0.089 |
| College and above | 2389 (71.9%) | 8823 (56.5%) |  |  |
| **Household annual income** |  |  |  |  |
| <35k or unknown | 788 (23.7%) | 5111 (32.7%) | 0.248 | 0.073 |
| 35-100k | 1420 (42.8%) | 6756 (43.3%) |  |  |
| >100k | 1113 (33.5%) | 3742 (24.0%) |  |  |
| **Health Insurance Status** |  |  |  |  |
| No | 223 (6.7%) | 2024 (13.0%) | 0.187 | 0.075 |
| Yes | 3098 (93.3%) | 13585 (87.0%) |  |  |
| **Food insecure** |  |  |  |  |
| No | 2801 (84.3%) | 11286 (72.3%) | 0.297 | 0.088 |
| Yes | 520 (15.7%) | 4323 (27.7%) |  |  |
| **Housing Instable** |  |  |  |  |
| No | 3028 (91.2%) | 13038 (83.5%) | 0.233 | 0.047 |
| Yes | 293 (8.8%) | 2571 (16.5%) |  |  |
| **Region** |  |  |  |  |
| MidWest | 590 (17.8%) | 2664 (17.1%) | 0.182 | 0.045 |
| NorthEast | 1104 (33.2%) | 4257 (27.3%) |  |  |
| South | 764 (23.0%) | 4741 (30.4%) |  |  |
| West | 832 (25.1%) | 3767 (24.1%) |  |  |
| US Terr or Unknown | 31 (1.0%) | 180 (1.2%) |  |  |
| **Access to primary care doctor** |  |  |  |  |
| No | 744 (22.4%) | 4303 (27.6%) | 0.158 | 0.102 |
| Yes | 2531 (76.2%) | 10672 (68.4%) |  |  |
| Don't know | 46 (1.4%) | 634 (4.1%) |  |  |
| **Current Smoker Status** |  |  |  |  |
| No | 2806 (84.5%) | 11637 (74.6%) | 0.248 | 0.087 |
| Yes | 515 (15.5%) | 3972 (25.4%) |  |  |
| **Obesity Status** |  |  |  |  |
| No | 2367 (71.3%) | 10807 (69.2%) | 0.045 | 0.003 |
| Yes | 954 (28.7%) | 4802 (30.8%) |  |  |
| **Cancer** |  |  |  |  |
| No | 3175 (95.6%) | 15086 (96.6%) | 0.054 | 0.059 |
| Yes | 146 (4.4%) | 523 (3.4%) |  |  |
| **Kidney disease** |  |  |  |  |
| No | 3267 (98.4%) | 15371 (98.5%) | 0.008 | 0.025 |
| Yes | 54 (1.6%) | 238 (1.5%) |  |  |
| **Lung disease** |  |  |  |  |
| No | 3245 (97.7%) | 15140 (97.0%) | 0.045 | 0.001 |
| Yes | 76 (2.3%) | 469 (3.0%) |  |  |
| **Type-2 diabetes** |  |  |  |  |
| No | 3086 (92.9%) | 14445 (92.5%) | 0.015 | 0.003 |
| Yes | 235 (7.1%) | 1164 (7.5%) |  |  |
| **Heart diseases** |  |  |  |  |
| No | 3241 (97.6%) | 15144 (97.0%) | 0.035 | 0.016 |
| Yes | 80 (2.4%) | 465 (3.0%) |  |  |
| **HIV/AIDS** |  |  |  |  |
| No | 3188 (96.0%) | 14917 (95.6%) | 0.021 | 0.025 |
| Yes | 133 (4.0%) | 692 (4.4%) |  |  |
| **Mental health disorders** |  |  |  |  |
| No | 2185 (65.8%) | 9514 (61.0%) | 0.101 | 0.048 |
| Yes | 1136 (34.2%) | 6095 (39.0%) |  |  |
| **Immunosuppression** |  |  |  |  |
| No | 3227 (97.2%) | 15280 (97.9%) | 0.047 | 0.064 |
| Yes | 94 (2.8%) | 329 (2.1%) |  |  |

377 \*SMD: standard mean difference

In the per-protocol (PP) analysis, the 6-month weighted incidence of long COVID, as defined by at least one symptom persisting for three months post-infection and concurrent activity limitation which were absent before infection, was 0.9% (95% CI: 0.5%, 1.3%) in the group who received a third COVID-19 vaccine dose and 1.0% (95% CI: 0.8%, 1.1%) in the group who did not receive a third dose, with an incidence difference of –0.1% (95% CI: -0.5%, 0.4%) and an incidence ratio of 0.929 (95% CI: 0.544, 1.437) (Table 2). A similar pattern was observed for other case definitions, such as symptoms persisting for three months at two time points, where the standardized incidence was 1.2% (95% CI: 1.0%, 1.4%) in the group who received a third COVID-19 vaccine dose and 1.3% (95% CI: 0.8%, 1.8%) in the group who did not receive a third dose, yielding a incidence difference of 0% (95% CI: -0.4%, 0.6%) and a incidence ratio of 1.032 (95% CI: 0.659, 1.577) (Supplementary Table 2A).

**Table 2:** Analog of the Per-Protocol IPTW and IPCW weighted incidence, incidence difference, and incidence ratio (95% confidence intervals) for 6- and 12-month long COVID, The CHASING COVID Cohort, September 2021 – December 2023

| Follow up time | Treatment Strategy | Weighted risk (95% CI) |  | Weighted Risk difference (95% CI) |  | Weighted Risk ratio (95% CI) |  |
| --- | --- | --- | --- | --- | --- | --- | --- |
| 6 months | Participants who received a third COVID-19 vaccine dose | 0.9% | 0.5%, 1.3% | -0.1% | -0.5%, 0.4% | 0.929 | 0.544, 1.437 |
|  | Participants who did not receive a third COVID-19 vaccine dose | 1.0% | 0.8%, 1.1% | ref | ref | ref | ref |
| 12 months | Participants who received a third COVID-19 vaccine dose | 4.9% | 4.1%, 5.9% | 0.4% | -0.5%, 1.4% | 1.092 | 0.897, 1.329 |
|  | Participants who did not receive a third COVID-19 vaccine dose | 4.5% | 4.1%, 4.8% | ref | ref | Ref | ref |

\* Long COVID was defined as the occurrence of $\geq 1$ new symptom and concurrent activity limitation between 3 – 12 months post-infection, both of which were absent in the year preceding infection.

For the 12-month PP analysis, individuals who received a third COVID-19 vaccine dose had an incidence of 4.9% (95% CI: 4.1%, 5.9%) compared to 4.5% (95% CI: 4.1%, 4.8%) in the group who did not receive a third dose, with an incidence difference of 0.4% (95% CI: -0.5%, 1.4%) and an incidence ratio of 1.092 (95% CI: 0.897, 1.329) (Table 2). For other definitions, such as symptoms persisting twice at three months, the incidence difference was 0.9% (95% CI: -0.1%, 2.1%) and the incidence ratio was 1.184 (95% CI: 0.989, 1.390) (Supplementary Table 2B). Cumulative incidence curves showed overlapping trends for the groups who received a third COVID-19 vaccine dose and the group who did not receive a third dose (Figure 1).

**Figure 1:**
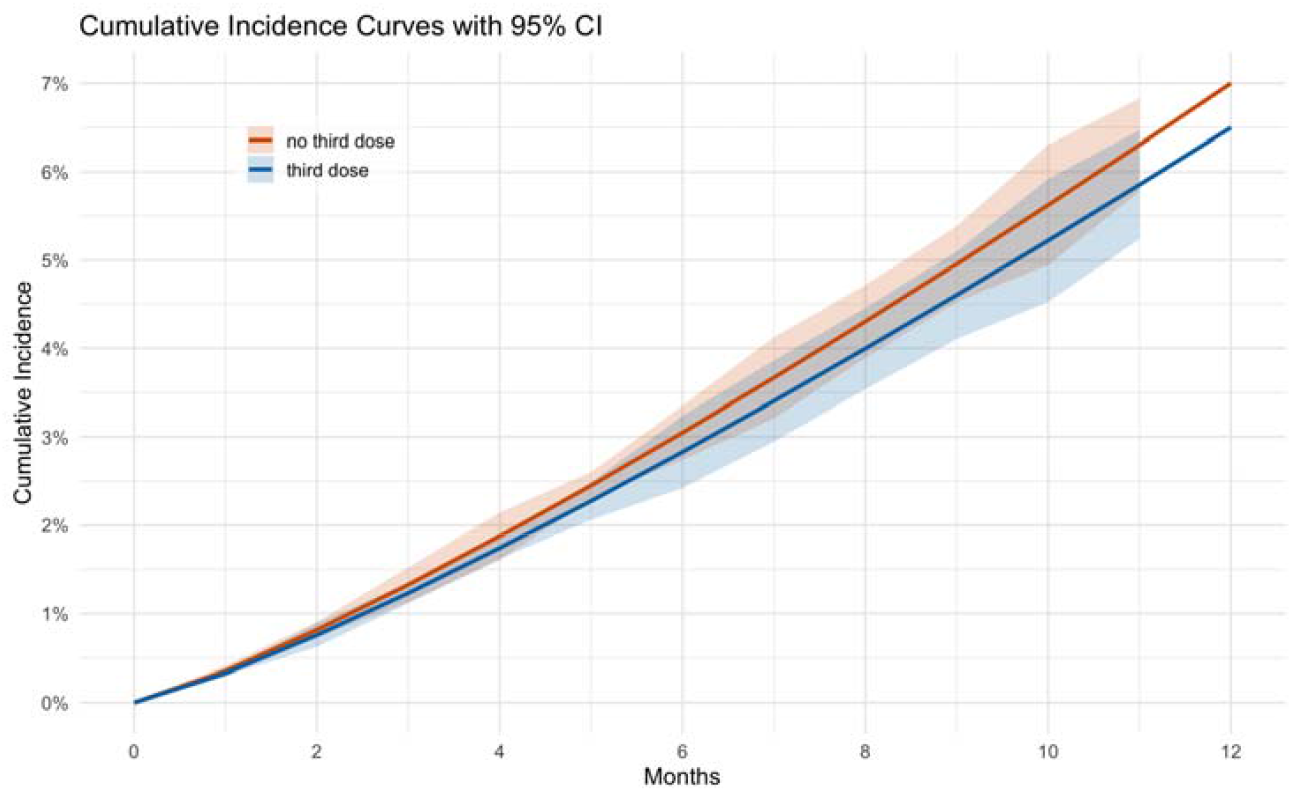
Estimated cumulative incidence of long COVID curves by the third dose vaccination status across 16 monthly sequential target trials between September 2021 and December 2023

## DISCUSSION

This study investigated the relationship between receiving a third dose of the COVID-19 vaccine (booster dose) and the 6- and 12-month incidence of long COVID among a U.S. community-based cohort. Our results showed no meaningful differences in the incidences of long COVID between the group who received at least a third COVID-19 vaccine dose and the group who did not in sequential prospective target trials. Notably, the null studies and our analysis tend to share designs that capture milder or community-reported infections and rely on participant-reported outcomes, whereas studies reporting a protective effect more often used electronic health records or claims data, which may preferentially sample individuals with more severe illness or healthcare contact. Thus, differences in case-ascertainment—particularly whether individuals with milder long COVID are included—may partly explain why some investigations observe an effect and others do not.

These findings contrast with several studies that suggested booster vaccination reduces risk of long COVID. The discrepancies can be attributed to several factors. Firstly, differences in study design, such as the cross-sectional approaches used by Domènech-Montoliu et al. and Romeiser et al. compared to our target trial emulation within a prospective community-based cohort, may explain the inconsistencies ^13,14^. Secondly, variability in long COVID definitions, ranging from persistent symptoms (Xie et al.) to broader measures including activity limitations (Di Fusco et al.), further complicates comparisons ^15,16^. We addressed this by testing multiple case definitions in sensitivity analyses, including alternative measures of symptom persistence and functional impact. Thirdly, behavioral factors, such as healthcare utilization disparities noted by Nguyen et al. and differences in vaccine types and schedules, as highlighted by Wee et al., may also contribute ^18,19^. These complexities emphasize the need for harmonized definitions and methodologies in future research.

A key strength of this study is the use of TTE design, which enhances causal inference by aligning observational design with the framework of RCT. Unlike standard approaches, TTE explicitly defines treatment strategies and time zero in advance, ensuring that eligibility, exposure classification, and follow-up begin simultaneously ^47^. This structure helps prevent immortal time bias—a form of bias that arises when participants must survive a certain period to receive the exposure, thus artificially lowering event rates in the exposed group ^48^. By emulating a sequence of trials over time, our design not only mitigate this bias but also improves statistical efficiency by incorporating all eligible person-time.

Despite its strengths, this study has limitations. First, long COVID outcomes relied on self-reported symptoms and conditions, which may introduce recall and reporting biases. Second, our sample excluded individuals with prior SARS-CoV-2 infections within three months of time zero, potentially limiting generalizability to populations with differing exposure histories. However, this approach aligns with CDC guidance, which suggests delaying COVID-19 vaccination for up to three months after infection, as recent infection offers some immunity ^49^. This strategy also helps reduce immortal time bias by excluding individuals who may have been temporarily at lower risk of reinfection during that period. Third, although our TTE design adjusted for a comprehensive set of confounders, unmeasured factors, such as health literacy differences between staying up-to-date vaccinated individuals and vaccinated once individuals, may still confound the results. Lastly, infection imputation algorithms—while designed to enhance detection of infections using repeated serologic and self-report data, may still be susceptible to misclassification. To reduce this risk, we applied conservative criteria when assigning infection dates; for instance, serology-identified infections were only assigned dates when supported by viral test results or Council of State and Territorial Epidemiologists criteria that helped narrow the plausible infection window. Nonetheless, some residual misclassification may persist due to limitations in accurately pinpointing the true infection date. Therefore, these limitations should be considered when interpreting the study’s findings.

Our findings suggested that receiving at least a third dose of the COVID-19 vaccine may not substantially reduce the risk of long COVID compared to only receiving the primary vaccine in a community-based cohort. These results underscored the need for further research employing rigorous analytical methods to clarify the relationship between vaccination and long COVID. Future studies could potentially benefit from prospective designs that include physician-diagnosed long COVID outcomes to reduce the risk of outcome misclassification and better collect potential differences (e.g. healthcare-seeking behavior and health literacy differences) between individuals who are up-to-date on COVID-19 vaccination and those who received only the primary vaccines.

## FUNDING

Funding for this project is provided by The National Institute of Allergy and Infectious Diseases (NIAID), award number UH3AI133675 (MPIs: D Nash and C Grov), Pfizer Inc, the CUNY Institute for Implementation Science in Population Health (cunyisph.org) and the COVID-19 Grant Program of the CUNY Graduate School of Public Health and Health Policy.

The NIH played no role in the production of this manuscript nor necessarily endorses the findings. The study design was developed by CUNY without input from Pfizer.

## PATIENT CONSENT STATEMENT

Informed consent forms were completed in a web browser on participants’ computer or mobile device at baseline, each round of serological testing, and at periodic follow-up assessments. The study was approved by the Institutional Review Boards of the City University Of New York (CUNY) Graduate School of Public Health and Health Policy (New York, NY, U.S.) (protocol 2020-0256).

## ACKNOWLEDGEMENTS

The authors wish to thank the participants of the CHASING COVID Cohort Study. We are grateful to you for your contributions to the advancement of science around the SARS-CoV-2 pandemic.

## AUTHOR CONTRIBUTORS

YS: Conceptualization; Study design; Data curation; Formal analysis; Investigation; Visualization; Writing—original draft; Writing—review & editing. DN: Conceptualization; Supervision; Resources; Funding acquisition; Project administration; Writing—review & editing. ZS: Methodology; Interpretation; Writing—review & editing. MR: Methodology; Interpretation; Writing—review & editing. KG: Methodology; Interpretation; Writing—review & editing.

## DATA AVAILABILITY

The data underlying this study contain potentially identifiable health information and are not publicly available due to ethical and legal restrictions. De-identified data may be shared on reasonable request and subject to institutional approvals and a Data Use Agreement. Requests will be reviewed case-by-case by the study investigators and the CUNY ISPH data governance/IRB. Please contact the corresponding author.

