## Supplementary Files for "Effect of a Third COVID-19 Vaccine Dose on the Incidence of Long COVID Among Adults Who Completed a Primary Vaccine Series: a Target Trial Emulation in a Community-Based Cohort"

### Appendix Table 1: STROBE (Strengthening the Reporting of Observational studies with Epidemiology) Checklist of items that should be included in reports of cohort studies.

|  | Item | Recommendation | Page |
| --- | --- | --- | --- |
| Title and abstract | 1 | (a) Indicate the study’s design with a commonly used term in the title or the abstract |  |
|  |  | (b) Provide in the abstract an informative and balanced summary of what was done and what was found | 1-2 |
| Introduction | | | |
| Background/rationale | 2 | Explain the scientific background and rationale for the investigation being reported | 3 |
| Objectives | 3 | State specific objectives, including any prespecified hypotheses | 3 |
| Methods | | | |
| Study design | 4 | Present key elements of study design early in the paper | 4 |
| Setting | 5 | Describe the setting, locations, and relevant dates, including periods of recruitment, exposure, follow-up, and data collection | 4 |
| Participants | 6 | (a) Give the eligibility criteria, and the sources and methods of selection of participants. Describe methods of follow-up | 5, Supp Table 1 |
|  |  | (b) For matched studies, give matching criteria and number of exposed and unexposed |  |
| Variables | 7 | Clearly define all outcomes, exposures, predictors, potential confounders, and effect modifiers. Give diagnostic criteria, if applicable | 4-5, Appendix 1, 2 |
| Data sources/ measurement | 8 | For each variable of interest, give sources of data and details of methods of assessment (measurement). Describe comparability of assessment methods if there is more than one group | 4-5, Appendix 1, 2 |
| Bias | 9 | Describe any efforts to address potential sources of bias | 4-5, Appendix 1, 2 |
| Study size | 10 | Explain how the study size was arrived at | Supp Figure 1&2 |
| Quantitative variables | 11 | Explain how quantitative variables were handled in the analyses. If applicable, describe which groupings were chosen and why | 4-5, Appendix 1, 2 |
| Statistical methods | 12 | (a) Describe all statistical methods, including those used to control for confounding |  |
|  |  | (b) Describe any methods used to examine subgroups and interactions | 6-7 |
|  |  | (c) Explain how missing data were addressed |  |
|  |  | (d) If applicable, explain how loss to follow-up was addressed |  |
|  |  | (e) Describe any sensitivity analyses |  |
| Results | | |  |
| Participants | 13 | (a) Report numbers of individuals at each stage of study—eg numbers potentially eligible, examined for eligibility, confirmed eligible, included in the study, completing follow-up, and analyzed | 7, Supp Figure 1&2 |
|  |  | (b) Give reasons for non-participation at each stage |  |
|  |  | (c) Consider use of a flow diagram |  |
| Descriptive data | 14 | (a) Give characteristics of study participants (eg demographic, clinical, social) and information on exposures and potential confounders | 7, Table 1 |
|  |  | (b) Indicate number of participants with missing data for each variable of interest |  |
|  |  | (c) Summarize follow-up time (eg, average and total amount) |  |
| Outcome data | 15 | Report numbers of outcome events or summary measures over time | 7-8, Table 2 |
| Main results | 16 | (a) Give unadjusted estimates and, if applicable, confounder-adjusted estimates and their precision (eg, 95% confidence interval). Make clear which confounders were adjusted for and why they were included  (b) Report category boundaries when continuous variables were categorized  (c) If relevant, consider translating estimates of relative risk into absolute risk for a meaningful time period | 7-8, Table 2 |
| Other analyses | 17 | Report other analyses done—eg analyses of subgroups and interactions, and sensitivity analyses | 7-8, Supp Table 2-3 |
| Discussion |  |  |  |
| Key results | 18 | Summarise key results with reference to study objectives | 7-8 |
| Limitations | 19 | Discuss limitations of the study, taking into account sources of potential bias or imprecision. Discuss both direction and magnitude of any potential bias | 9 |
| Interpretation | 20 | Give a cautious overall interpretation of results considering objectives, limitations, multiplicity of analyses, results from similar studies, and other relevant evidence | 8-9 |
| Generalisability | 21 | Discuss the generalisability (external validity) of the study results | 9 |
| Other information |  |  |  |
| Funding | 22 | Give the source of funding and the role of the funders for the present study and, if applicable, for the original study on which the present article is based | 10 |

### Appendix 1: SARS-CoV-2 Infection and Dates

Infection status was classified based on one or more of the following criteria: (1) a self-reported positive result from a polymerase chain reaction (PCR) or rapid antigen test, regardless of whether it was administered by a healthcare provider or completed at home; (2) serologic evidence of prior infection, indicated by the detection of total antibodies (IgA, IgM, or IgG) against the SARS-CoV-2 nucleocapsid protein from dried blood spot (DBS) specimens collected through the study; or (3) classification as a probable case according to the Council of State and Territorial Epidemiologists (CSTE) definition. Infection dates were determined by a self-reported exact date or were imputed based on the earliest and latest plausible dates of infection—creating an infection window bounded by the date of survey collection, reported symptoms and epidemiologic linkage, or serologic sample collection, depending on the infection classification method.

(1) Infection Identified by Positive PCR or Antigen Tests: Confirmed SARS-CoV-2 infection was determined through self-reported positive viral PCR or antigen test, either conducted by a healthcare provider or exclusively using a home-based rapid test. The infection dates were primarily based on a self-reported infection date. If participants provided only the month without a specific day, the midpoint of that month was assigned as the infection date. The infection date was imputed as the midpoint between the date they reported the infection and the previous follow-up assessment, reflecting the question about positive viral tests since the last survey.

(2) Infections Identified by Positive Serology Tests: We also identified confirmed SARS-CoV-2 infections using serologic testing results from study-collected dried blood spots (DBS) at 4 time points: April-September 2020 (Serology Period 1), November 2020-March 2021 (Serology Period 2), March-June 2022 (Serology Period 3), and July 2024-October 2024 (Serology Period 4). For serology-identified infections accompanied by a self-reported positive PCR or antigen test, or meeting CSTE criteria, infection dates were determined using the imputation intervals described in the relevant sections. Serology-identified infections without a prior viral or serology test or CSTE evidence were excluded, as infection dates could not be reliably estimated within a 90-day interval.

(3) Infections Identified by CSTE Criteria: Given the limited testing capacity during the initial surge of Omicron infections in the U.S. ^1–4^, we identified probable infections occurring between December 6, 2021 and January 11, 2022 using the Council of State and Territorial Epidemiologists (CSTE) probable case definition ^5^. Participants reported: 1) experiencing either at least one of the following symptoms — cough, shortness of breath, loss or altered sense of smell, loss or altered sense of taste, or chest pain; or at least two of the following symptoms — fever, chills, myalgia, headache, sore throat, nausea/vomiting, congestion/runny nose; and 2) having epidemiologic linkage in the prior 14 days, including close contact with a confirmed or probable SARS-CoV-2 case or being a healthcare worker. Since the epidemiologic linkage was assessed as two weeks prior to the follow-up assessment, the infection date was assigned as the midpoint between the assessment date and 14 days prior.

### Appendix 2. Supplementary Methods for Time-Fixed and Time-Updated Characteristics

Time-fixed characteristics

Time-fixed characteristics—including age, gender, race/ethnicity, education, household income, smoking status, obesity, and chronic conditions—were collected between March and July 2020. Participants who reported smoking cigarettes or using electronic products every day or on some days were considered smokers. Obesity was defined as a BMI ≥30 kg/m² ^6^. Participants were asked to report pre-existing conditions identified by CDC as risk factors for severe COVID-19 by responding to the prompt, “Please select all that apply,” which included current asthma, cardiovascular disease (heart attack, angina, coronary heart disease), high blood pressure, cancer, chronic kidney or liver disease, chronic lung disease (asthma, COPD), type 2 diabetes, HIV/AIDS, immunosuppression, and mental health conditions (depression, PTSD, or anxiety) ^7^. All chronic conditions were coded as binary indicators.

Time-updated characteristics

Between July 2020 and December 2023, participants completed follow-up assessments approximately every 3-6 months, reporting on health insurance coverage, access to a primary care doctor, region of residence, food security, and housing stability. Health insurance coverage was assessed using a question adapted from the 2014 Behavioral Risk Factor Surveillance System (BRFSS) survey ^8^. Participants were classified as uninsured if they responded “no” or “don’t know” to whether they had any form of health insurance, including employer-sponsored insurance, prepaid plans, or government program. Health insurance and access to a primary care doctor were categorized as “Yes,” “No,” or “Don’t know,”. Region of residence was determined based on ZIP code reported in each follow-up. Food insecurity was assessed using questions from the U.S. Department of Agriculture (USDA) Household Food Security Survey (HFSS) ^9^. Participants responded, “often true”, “sometimes true”, or “never true” to three statements: (1) “We couldn’t afford to eat balanced meals”, (2) “We worried whether our food would run out before we got money to buy more”, and (3) “The food that we bought just didn’t last, and we didn’t have money to get more”. Those answering “often true” or “sometimes true” to any statements were classified as food insecure ^10^. Housing instability was assessed using a BRFSS-adapted question on how often participants worried about affording rent or mortgage payments ^11^. Responses of “Always” or “Usually” indicated housing instability, while “Sometime”, “Rarely”, and “Never” indicated housing stability ^12^.

Supplementary Figure 1: Flowchart of Participant Selection and Eligibility Criteria for the First Sequential Trial (September 2021) in the CHASING COVID Cohort Study

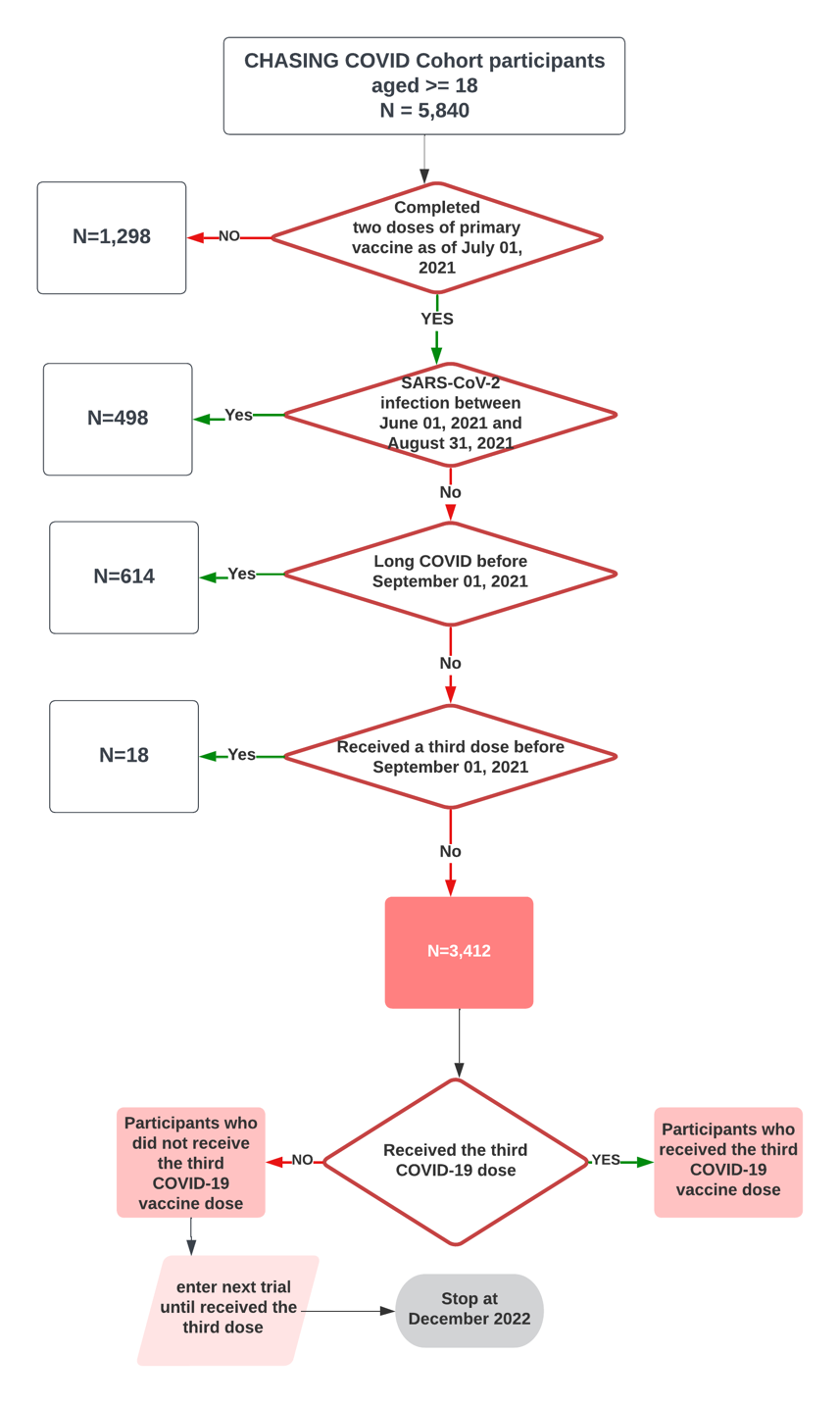

Supplementary Figure 2: Sequential trial assembly and the determination of time zero for each trial

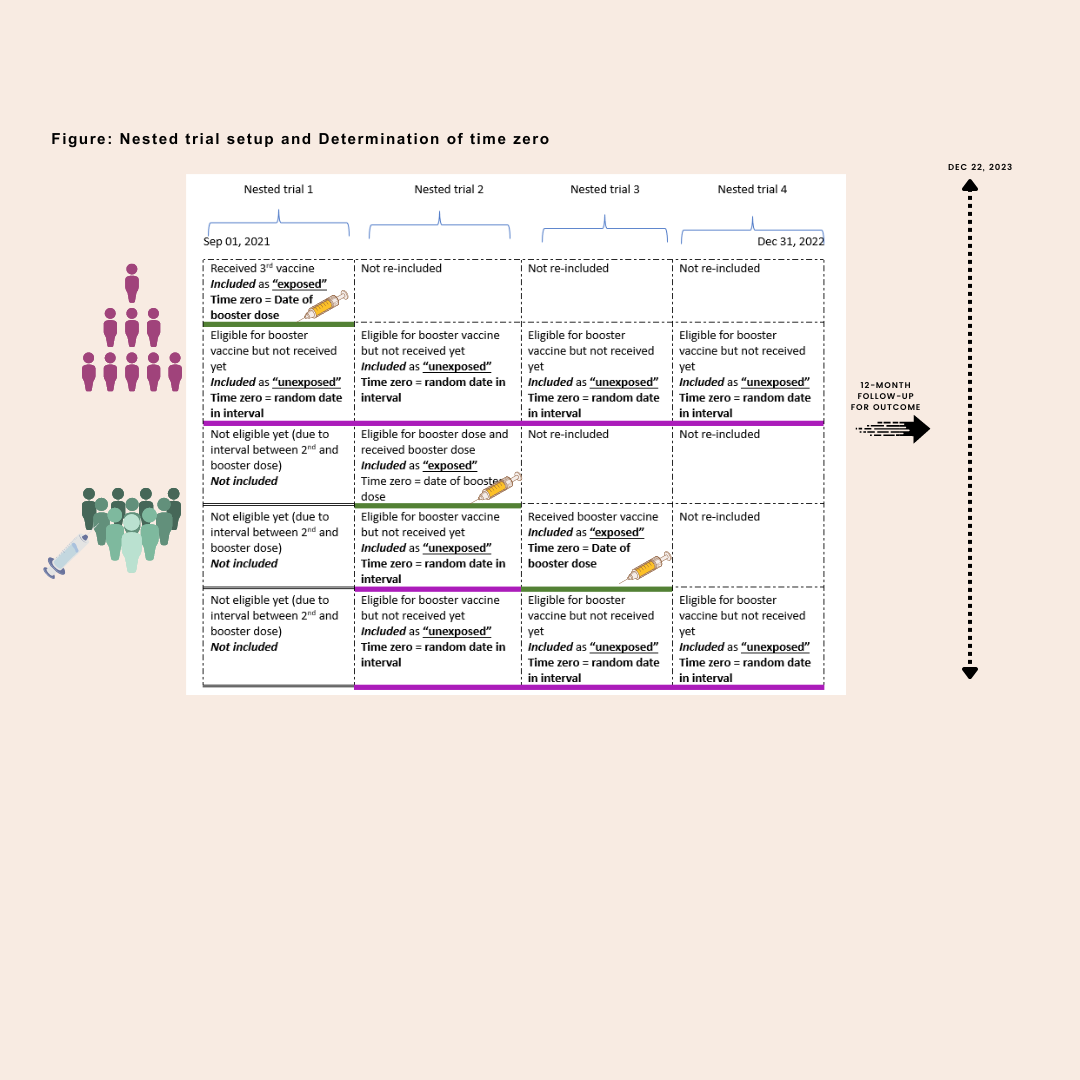

Note: In this hypothetical example with only four 1-month intervals, there are five total individual participants assessed. Three patients (green-filled outlines on the bottom) received a vaccine, and two patients (purple-filled outlines on the bottom) received no vaccine dose during four 1-month blocks. There are a total of 8 emulated participants in the unexposed arm and 3 emulated participants in the exposed arm, because participants can be included as unexposed participants until the month of an vaccine receipt.

### Supplementary Table 1: Specification and emulation of a target trial of the third dose of COVID-19 vaccine and 6- and 12-month Long COVID risk among CHASING COVID Cohort study adult participants in the U.S., September 2021-December 2023

| Protocol | Target Trial Specification | Target Trial Emulation: Cohort design |
| --- | --- | --- |
| Eligibility Criteria | Inclusion criteria:  · Include individuals 18+ years of age  · Include participants who have received both doses of the two-dose primary vaccine series (Pfizer, Moderna, Novavax)  Exclusion criteria:  · Exclude participants who have a history of Long COVID  · Exclude participants who had a prior SARS-CoV-2 infection in the prior three months  · Exclude participants who had received any dose of COVID-19 dose beyond the primary series | Same as the target trial |
| Treatment Strategies | Participants will be assigned to:  1) the third dose of COVID-19 vaccine between September 1^st^, 2021 and December 31^st^, 2022 2) No third dose of COVID-19 vaccine | Same as the target trial |
| Treatment Assignment | 1) Individuals are randomly assigned to a strategy at time zero 2) Study is not blinded to investigators or enrollees | To emulate randomization by inverse probability of weighting |
| Outcomes | Long COVID was defined as the presence of at least one symptom occurring at or beyond three months post-infection, accompanied by concurrent limitations in daily activities, with both not reported in the year prior to infection. | Same as the target trial |
| Follow-up | In each nested trial, follow-up begins at date of assignment and continues until earliest of 1) development of the outcome of interest (Long COVID) 2) loss to follow-up for any follow-up assessment post-time zero  3) administrative end of 6 or 12 months follow-up after time zero | Same as the target trial |
| Causal Contrasts | Intention-to-treat effect (ITT)  Per-protocol effect (PP) | Observational analogue of ITT effect and PP effect |
| Statistical Analysis | We will apply inverse probability censoring weights (IPCW) of pre- and post-time zero prognostic factors associated with loss to follow-up and deviation of treatment assignment. | Same as for the target trial with adjustment for IPTW of factors in the observational analogue of treatment assignment |

### Supplementary Table 2A: Analog of the per-protocol weighted risk, risk difference, and risk ratio (95% confidence intervals) for 6-month Long COVID, The CHASING COVID Cohort, September 2021 – December 2023

| Long COVID case definitions | Treatment Strategy | Weighted Risk (95% CI) | | Weighted Risk difference  (95% CI) | | Weighted  Risk ratio  (95% CI) | |
| --- | --- | --- | --- | --- | --- | --- | --- |
| Symptom once ≥3 months post infection | Participants who received a third COVID-19 vaccine dose | 1.5% | 1.1%, 2.1% | 0% | -0.5%, 0.6% | 1.020 | 0.681, 1.454 |
|  | Participants who did not receive a third COVID-19 vaccine dose | 1.5% | 1.3%, 1.7% | Ref | ref | ref | ref |
| Symptom twice ≥3 months post infection | Participants who received a third COVID-19 vaccine dose | 1.3% | 0.8%, 1.8% | 0% | -0.4%, 0.6% | 1.032 | 0.659, 1.577 |
|  | Participants who did not receive a third COVID-19 vaccine dose | 1.2% | 1.0%, 1.4% | ref | ref | Ref | ref |
| Symptom once + self-identified Long COVID | Participants who received a third COVID-19 vaccine dose | 0.3% | 0.1%, 0.6% | 0% | -0.2%, 0.4% | 1.123 | 0.319, 2.432 |
|  | Participants who did not receive a third COVID-19 vaccine dose | 0.3% | 0.2%, 0.4% | ref | ref | Ref | Ref |
| Symptom once + limitation in daily activity | Participants who received a third COVID-19 vaccine dose | 0.9% | 0.5%, 1.3% | -0.1% | -0.5%, 0.4% | 0.929 | 0.544, 1.437 |
|  | Participants who did not receive a third COVID-19 vaccine dose | 1.0% | 0.8%, 1.1% | ref | ref | ref | ref |

### Supplementary Table 2B: Analog of the per-protocol weighted risk, risk difference, and risk ratio (95% confidence intervals) for 12-month Long COVID, The CHASING COVID Cohort, September 2021 – December 2023

| Long COVID case definitions | Treatment Strategy | Weighted Risk (95% CI) | | Weighted Risk difference  (95% CI) | | Weighted  Risk ratio  (95% CI) | |
| --- | --- | --- | --- | --- | --- | --- | --- |
| Symptom once ≥3 months post infection | Participants who received a third COVID-19 vaccine dose | 7.2% | 6.2%, 8.2% | 0.6% | -0.4%, 1.8% | 1.101 | 0.934, 1.284 |
|  | Participants who did not receive a third COVID-19 vaccine dose | 6.5% | 6.1%, 7.0% | Ref | ref | Ref | ref |
| Symptom twice ≥3 months post infection | Participants who received a third COVID-19 vaccine dose | 6.3% | 5.5%, 7.4% | 0.9% | -0.1%, 2.1% | 1.184 | 0.989, 1.390 |
|  | Participants who did not receive a third COVID-19 vaccine dose | 5.4% | 5.0%, 5.8% | ref | ref | Ref | Ref |
| Symptom once + self-identified Long COVID | Participants who received a third COVID-19 vaccine dose | 1.5% | 1.0%, 2.0% | 0.4% | -0.2%, 0.9% | 1.358 | 0.858, 1.930 |
|  | Participants who did not receive a third COVID-19 vaccine dose | 1.1% | 0.9%, 1.3% | ref | ref | Ref | Ref |
| Symptom once + limitation in daily activity | Participants who received a third COVID-19 vaccine dose | 4.9% | 4.1%, 5.9% | 0.4% | -0.5%, 1.4% | 1.092 | 0.897, 1.329 |
|  | Participants who did not receive a third COVID-19 vaccine dose | 4.5% | 4.1%, 4.8% | ref | ref | Ref | ref |

### Supplementary Table 3: Analog of the intention-to-treat weighted incidence, incidence difference, and incidence ratio (95% confidence intervals) for 6- and 12-month Long COVID, the infection naïve individuals, The CHASING COVID Cohort, September 2021 – December 2023

| Follow up time | Treatment Strategy | Weighted Risk (95% CI) | | Weighted Risk difference  (95% CI) | | Weighted  Risk ratio  (95% CI) | |
| --- | --- | --- | --- | --- | --- | --- | --- |
| 6 months | Participants who received a third COVID-19 vaccine dose | 0.9% | 0.6%, 1.8% | -0.1% | -0.4%, 0.4% | 0.921 | 0.634, 1.478 |
|  | Participants who did not receive a third COVID-19 vaccine dose | 1.0% | 0.5%, 1.5% | ref | ref | ref | ref |
| 12 months | Participants who received a third COVID-19 vaccine dose | 4.6% | 2.4%, 6.1% | 0.4% | -1.1%, 1.7% | 1.107 | 0.902, 1.358 |
|  | Participants who did not receive a third COVID-19 vaccine dose | 4.2% | 3.0%, 5.7% | ref | ref | ref | ref |
